# Assessing the cross-cultural applicability of a mental health literacy questionnaire for young people in rural Malawi

**DOI:** 10.64898/2025.12.07.25341223

**Authors:** Cameron Goldsmith, Joel Nyali, Sandra Jumbe

## Abstract

In Malawi, mental health resources remain limited while demand continues to rise, particularly among young people, resulting in a substantial treatment gap. Enhancing mental health literacy (MHL) is increasingly recognised as a preventive strategy to reduce the onset and escalation of mental disorders. MHL is commonly assessed through instruments evaluating knowledge, beliefs, self-help strategies, and help-seeking behaviours. Although previous studies in Malawi have explored mental health awareness, none have directly measured MHL among young people, and few have addressed the cultural adaptations necessary for contextual validity. Such adaptations are essential to support decolonising research practices and to generate accurate insights into local understandings of mental health.

This explanatory mixed-methods study examined the applicability of a Portuguese-developed Mental Health Literacy Questionnaire (MHLq) among young people in rural Malawi. Quantitative data (n = 107) were statistically analysed, followed by qualitative focus groups (n = 7) to interpret and contextualise findings. The dataset yielded moderately high mean MHL scores, contrasting with participants’ perceptions that mental health literacy in Malawi remains low. Thematic analysis identified three central themes: (1) epistemic exclusion, (2) conceptual misfits, and (3) translation challenges.

Findings highlight the need to adapt the MHLq to incorporate culturally relevant knowledge, linguistic nuance, and local conceptualisations of mental health. Avoiding psychiatric terminology that lacks direct Chichewa equivalents is particularly important. The study underscores the necessity of participatory, contextually grounded approaches for developing and validating MHL tools in low-resource, culturally diverse settings.

## Background

### Mental health in Malawi

Mental health is becoming an increasingly significant issue in Malawi, particularly among young people, where there is a 10-30% prevalence of common mental health conditions and a continuously rising suicide rate for this age group (Udedi et al 2013; Banda et al., 2021)). However, Malawi’s mental healthcare system suffers from a severe lack of resources with just three mental health units, located in urban areas and staffed by 3-4 psychiatrists nationwide and one psychiatric nurse per 100,000 population (Udedi et al., 2013). This is inadequate and unsustainable for a population of 21.1 million people, who primarily (80%) reside in rural areas (World Bank, 2023; Udedi et al., 2013).

It has been well established that individuals can be profoundly impacted by untreated mental health, resulting in poor outcomes in their social lives, physical health, and ability to perform day-to-day functions (Kaushik, Kostaki, & Kyriakopolos, 2016). Young people in Malawi are experiencing this impact, especially since the Covid-19 pandemic, which has exacerbated already existing issues such as deprivation, health challenges, and substance misuse, in addition to mental health issues (Jumbe et al., 2022). This necessitates a prevention approach to mental health that is both cost-effective and reduces pressures on Malawi’s under-resourced and overburdened system.

### Mental health literacy

Mental health literacy, which encapsulates knowledge and awareness about mental health, is an important concept for preventing mental health issues at a population level. There are five key components of mental health literacy, including knowledge about: preventing mental disorders, recognising mental disorders, self-help strategies, help-seeking and treatment options, and mental health first aid skills (Jorm, 2012). A focus on mental health literacy is a low cost and effective strategy for preventing mental health issues by increasing the uptake of early help options, reducing stigma, and empowering communities to support themselves and one another (Sequiera et al., 2022).

However, many existing mental health literacy tools are developed in high-income countries and are later adapted for use in low- and middle-income countries. This can undermine local knowledge and belief systems, such as traditional healing practices. Malawi has a long history of traditional mental health practices tied to local spiritual and religious beliefs. Traditional spiritual healers and religious leaders are widely respected within local communities (Steinforth, 2017). They address mental distress through various treatment tactics, including magic and herbal remedies (Read et al., 2009). Although it is common for people in Malawi to seek care from traditional healers, spiritual healers, or religious leaders before or instead of a mental health professionals (Udedi, 2016; Wright et al., 2014), traditional diagnoses are usually dismissed as incomprehensive and undescriptive of clinically treatable mental disorders (Steinforth, 2008; Steinforth, 2017).

Two previous pilot studies for mental health education programmes in Malawi determined that both student-centred (Kutcher et al., 2019) and educator-centred (Kutcher et al., 2015) mental health literacy programmes improve attitudes toward mental health. However, these programmes were originally developed and validated in Canada before being adapted and implemented in Malawi (Kutcher et al., 2015; Kutcher et al., 2019). Although some cultural adaptations were made, the tool fails to address Malawi’s traditional mental health practices, and rather, focuses primarily on psychiatric mental health education. These pitfalls in mental health literacy measurement tools and psychiatric mental healthcare in general signify a need for a more culturally appropriate approach to preventing mental disorders in Malawi and other low and middle income countries (Chibanda et al., 2021).

### Epistemic injustice in mental health literacy tools

To address this gap in the current literature, the epistemic injustice, the silencing of cultural beliefs and ideologies, that is committed when cultural contexts and ‘truths’ are ignored or dismissed, must be interrogated (Kidd, Medina, & Polhaus, 2017) (**Table 1**). In order to achieve this, psychiatric mental health must adopt a decolonial approach to improve mental health outcomes in postcolonial nations, such as Malawi. According to de Sousa Santo’s Epistemologies of the South, this might include recovering diverse knowledge systems, decolonizing research and history, and committing to anti-colonial practice (de Sousa Santos, 2014).

**Table 1.** Mental health epistemologies in Malawi. Two mental health epistemologies, psychiatric and traditional, exist in Malawi. However, the two do not exist harmoniously.

| **Western psychiatric healthcare** | **Traditional mental healthcare** |
| --- | --- |
| Scientific evidence-base | Spiritual and religious history |
| Derived from thinking in high income countries | Based on local traditions |
| English language | Local language |
| Individualistic approach | Community-based approach |
| Lacks cultural adaptations | Culturally relevant |

In this study, a material semiotics theoretical approach was first used to investigate how the two epistemologies, psychiatric and traditional mental health, interact within the MHLq and the responses it received from young Malawians. It was then used to propose suggestions for how to address the lack of access to decolonized and culturally appropriate mental health knowledge and literacy tools. Material semiotics, first coined by Michel Callon in 1982, is an approach to social analysis that dissects how social practices inform both discourse and its meaning (semiotics) and physical entities, such as bodies and technologies (material) (Law, 2019; Law, 2008). This rejects epistemic injustice by acknowledging that there is no single culture or social structure (Law, 2008). Instead, it focuses on what makes up the social fabric, how it came to be, what it includes and excludes from its practices, and how it interacts with other social fabrics and cultures (Law, 2008). The perpetuation of psychiatric mental health as the single “truth” in global mental health fails to achieve decolonization in countries like Malawi by dismissing the validity of traditional knowledge and treatment in the local context (Law, 2015). This, therefore, creates a binary between the two epistemologies, psychiatric and traditional mental health, where one is seen as valid and other as invalid in modern practice. As illustrated in Table 1, although these two epistemologies exist in Malawi, they do not exist harmoniously. A material semiotics approach challenges this by recognising that “realities are done in practice,” and therefore, multiple “truths” can exist simultaneously (Mol, Smith, & Weintraub, 2002). This is necessary in order to offer a decolonized and locally accepted mental health system and increase mental health literacy in Malawi in a way that is also meaningful to the psychiatric mental health field.

### Research Aims

This study used a material semiotics approach to assess a self-reporting mental health literacy questionnaire (MHLq) adapted from a prior study conducted in Portugal to assess mental health literacy in young people (Dias et al., 2018). Results from the Portuguese study demonstrated a mean total mental health literacy score of 105.27, with significant difference between respondents based on demographics, including sex (p<0.01) and proximity to people with mental health problems (p<0.01) (Dias et al., 2018). In this study, the MHLq was translated from English into Chichewa, the national language of Malawi alongside English, and distributed to respondents in rural Malawi (n= 107) to assess mental health literacy of young people, aged 15-35 years. No other cultural adaptations were made.

The first aim of this study was to assess the applicability of the MHLq for measuring young Malawians’ understandings and lived experiences of mental healthcare. The second aim was to recommend adaptations that better reflect the knowledge and lived experiences of young people in Malawi.

### Methodology

This study used an explanatory mixed methods research design, analysing quantitative MHLq data before conducting qualitative focus groups to explain the quantitative findings. This was done to identify MHLq responses that showed the most discrepancy between respondents, and explain the causes and consequences of those discrepancies through in-depth qualitative exploration. This aided in establishing the cultural applicability of this tool for use in Malawi.

### MHLq selection and translation

Details on the translation process along with preliminary validation and reliability results for this dataset were published in a separate paper (Jumbe, Nyali, & Newby, 2023). The full questionnaire can be found in **Supplementary Figure 1**.

### Chichewa MHLq participant recruitment

Research ethics approval was obtained from the National Committee on Research in the Social Sciences and Humanities (NCRSH) in Malawi on 16th March 2021, prior to commencing with the current study (Ref. No. P12/20/539).

Surveying was conducted by fieldworkers from Drug Fight Malawi and the National Youth Council of Malawi using convenience sampling between 25th March 2021 and 1st May 2021. The key aim for initially targeting rural communities was to pilot applicability of the Chichewa translated MHLq in this setting. The fieldworkers approached young people from four districts located in rural Malawi, Kasungu, Mchinji, Mzimba, and Salima.

Participants fulfilling the following inclusion criteria were recruited: young adults living in rural communities; aged 15 to 35 years. Exclusion criteria: not literate in Chichewa; unable to read and write independently. Data collection followed ethical guidelines with all participants providing written informed consent prior to completing the survey. Details of this process were published in a separate paper (Jumbe, Nyali, & Newby, 2023). Participants subsequently completed part one of the questionnaire (socio-demographics section), then part two which contained 29 MHLq items. Demographic characteristics of these 107 respondents are summarised in **Supplementary Table 1.**

### Focus group participant recruitment

The focus groups were conducted at a university in Blantyre, Malawi. Focus group participants were recruited using convenience sampling via an advert, posted on Twitter and university campus bulletin boards. The inclusion criteria were male or female, 18-35 years of age, and Malawi resident. Young people (n=11) contacted the research team by e-mail or WhatsApp to express their interest and give consent to participate. Relevant demographic information is summarised in **Table 4**. Informed consent was obtained through a consent form and a participant information sheet highlighting the study purpose, focus group session details, including mode of participation (online via Zoom or joining in person on the university campus), and how the data would be collected and used. This blended approach (both virtual and face-to-face) was conducted over a one-month period. All interested participants sent their signed consent forms for study participation and publication to the research team before the first focus group session.

### Data analysis

The MHLq consists of 29 items with categorical responses on a 5-point Likert scale, ranging from strongly disagree to strongly agree. Categorical responses are quantified by assigning each response with a number: 1= strongly disagree, 2= disagree, 3= neither agree nor disagree, 4= agree, and 5= strongly agree for all items, except #6, 10, 13, 15, 21, and 23 which were scored in reverse (Dias et al., 2018). MHLq items are then divided into four factors to quantify different aspects of mental health literacy **(Table 2).**

**Table 2.** Focus group sessions information.

| Session | Discussion topic | Session date | Session length (hr:min) |
| --- | --- | --- | --- |
| 1 | Factor 4: Self-help strategies<br>*1, 7, 19, 26 | 11 April 2022 | 00:59 |
| 2 | Factor 1: Knowledge of mental health problems<br>*2, 3, 9, 12, 16, 20, 22, 24, 25, 27, 28 | 14 April 2022 | 01:25 |
| 3 | Factor 2: Erroneous beliefs or stereotypes (part 1)<br>*6, 10, 11, 13, 14, 15, 21, 23 | 25 April 2022 | 01:40 |
| 4 | Factor 2: Erroneous beliefs or stereotypes (part 2) | 12 May 2022 | 01:34 |
|  | Factor 3: First aid skills and help-seeking behaviour<br>*4, 5, 8, 17, 18, 29 |  |  |
| * MHLq items that correspond with the four factors (Dias et al, 2018) indicating four different areas that make up mental health literacy |  |  |  |

In this study, scores for each factor were calculated and added together to generate each respondent’s total mental health literacy score with a potential range from a minimum score of 29 to a maximum score of 145 (**Supplementary Table 1**). An ordinary one-way ANOVA analysis with multiple comparisons was done using Microsoft Excel, to stratify mean total mental health literacy scores by demographic: age, sex, marital status, district of resident, education, occupation, and proximity to someone with a mental illness. Statistical significance was determined as *p<0.05.

Focus group sessions occurred in four parts, each session dedicated to discussing the MHLq items and concepts corresponding to each factor **(Table 2)**. A focus group discussion guide was prepared following quantitative analysis **(Table 3).** Prior to the first focus group session, participants were given two copies of the MHLq, one in Chichewa and one in English. At the start of the first session, participants were given an introductory presentation outlining key findings from the MHLq rural survey dataset. Each focus group session lasted approximately 60-90 minutes. Sessions were conducted primarily in English, though some Chichewa was spoken throughout.

**Table 3.** Focus group discussion guide. A focus group discussion guide was prepared following quantitative analysis of MHLq responses. The discussion guide highlighted themes and specific items from each MHLq Factor which require further explanation to understand the discrepancies between responses.

| Factor | Discussion points |
| --- | --- |
| 4 (self-help strategies) | □ Explore general thoughts on the self-help strategies identified in the MHLq<br>□ Explore what participants do to help their own |
|  | mental health |
| 1 (knowledge of mental health problems) | □ Explore general thoughts on the mental health knowledge tested in the MHLq<br>□ Explore participants understanding about mental health problems and conditions<br>□ Explore thoughts on the relevance and translation for items |
| 2 (erroneous beliefs/stereotypes) | □ Explore participants' general mental health attitudes<br>□ Explore the erroneous beliefs, stereotypes, and perceptions about mental health that participants are aware of<br>□ Explore general thoughts on the erroneous beliefs and stereotypes identified in the MHLq<br>□ Explore thoughts on the relevance and translation for items |
| 3 (first aid skills and help-seeking behaviours) | □ Explore participants' knowledge about different types of mental health professionals<br>□ Explore participants' access to mental health professionals<br>□ Explore thoughts on the relevance and translation for items<br>□ Explore the role of and access to traditional healers for mental health support |

Focus group sessions were recorded on Zoom and transcribed using Trint software. Recordings were listened to for transcript verification. Qualitative thematic analysis was performed to analyse the transcripts, initiated following the succession of the final focus group session in accordance with the Consolidated Criteria for Reporting Qualitative Research (Tong, Sainsbury, & Craig, 2007). First, transcripts were reread to identify commonalities within the text. Then, grounded theory was applied to the text, allowing commonalities to appear without testing a pre-existing hypothesis. Line-by-line analysis was used to code the transcripts to generate seven codes. Intersections between the codes were identified and codes were consolidated into three key themes: 1) conceptual misfits, 2) epistemic exclusion, and 3) translation challenges. Data saturation was reached after analysis of the transcript from the fourth session, and therefore, no further sessions were required.

### Theoretical framework

This study used a material semiotics approach to investigate the applicability of the MHLq for young people in Malawi. The goal of adopting this approach was to eliminate the hierarchy of social practices by denying the existence of a perceived dominant practice and culture while amplifying the validity of the perceived non-dominant practice and culture (Mol, Smith, & Weintraub, 2002). This does not disregard the materiality of any particular subject, but rather, highlights the meaning that it carries, as meaning greatly depends on positionality (Yates-Doerr, 2020; Haraway, 2016).

This approach is appropriate for analysing the cultural applicability of the MHLq for young people in Malawi. This is because questionnaires, in general, rely on metrics that apply statistics to and make generalisations about a given population (Yates-Doerr, 2020). Statistics and generalisation can be useful for establishing trends and assigning significance to those trends; however, the nuances of the social world often gets lost in such measurements (Law, 2015). A material semiotics approach to questionnaires goes beyond these metrics by investigating what the questionnaire does by looking at the impact of the questionnaire itself on both those doing the questionnaire and those responding to it (Yates-Doerr, 2020).

In this study, what the MHLq does and how it affects both mental health literacy and young people in Malawi are interrogated using this material semiotics approach. This shaped the focus group discussion guide, establishing where generalisations and trends could not be formed from MHLq responses and where nuance may have been lost in Likert scale measurements. It also shaped the use of grounded theory to allow the relationship between the MHLq and young Malawians to reveal itself during thematic analysis.

## Results

### Key findings from preliminary Chichewa MHLq responses

Preliminary MHLq data collected from 107 respondents in rural Malawi revealed total mental health literacy scores that were relatively higher than the Portuguese cohort. The mean total mental health literacy score was 114.63 (SD= 10.33) for the rural Malawi cohort **(Supplementary Table 2)** whereas the Portuguese cohort mean total score was 105.27 (SD= 7.05) (Dias et al., 2018). Therefore, suggesting that, on average, young people in Malawi’s rural community settings have relatively higher mental health literacy than young people in Portugal who were typically university students aged 18 to 25 years.

Ordinary one-way ANOVA with multiple comparisons statistical analysis did not reveal any significant difference in mean total mental health literacy score based on demographics with the Malawi cohort **(Table 5)**. This contrasted with results from the Portuguese cohort where significant differences in mean total mental health literacy scores were found based on sex (p<0.01) and proximity to someone with a mental illness (p<0.01) (Dias et al. (2018). These MHLq results were discussed in more detail during focus group sessions.

**Table 4.** Demographics of focus group participants.

| Participant (#) | Age | Sex (M/F) | Occupation | Field of study/ work |
| --- | --- | --- | --- | --- |
| 1 | 26-29 | Male | Employed | Research Assistant |
|  |  |  | Fulltime |  |
| 2 | 30-33 | Female | Student | Social Work |
| 3 | 22-25 | Male | Student | Social Work |
| 4 | 18-21 | Male | Student | Social Work |
| 5 | 26-29 | Male | Employed<br>Fulltime | Psychosocial<br>Counsellor |
| 6 | 22-25 | Male | Student | Banking and Finance |
| 7 | 22-25 | Male | Student | Business<br>Administration |

**Table 5.** Average total MHL scores stratified by demographic. MHLq responses were collected from young people in Malawi (n=107) and scored on a Likert scale. The table shows mean total MHL scores by demographic: age, sex, marital status, district of residence, education, occupation, and proximity to someone with a mental illness (mean ± SD, ordinary one-way ANOVA with multiple comparisons, *p< 0.05).

| Demographic | Count | Mean ± SD |
| --- | --- | --- |
| *Age* |  |  |
| 15-19 | 19 | 115.11 ±12.60 |
| 20-24 | 63 | 112.59 ±15.66 |
| 25-29 | 19 | 114.26 ±9.77 |
| 30-35 | 3 | 119.33 ±6.66 |
| Unknown | 2 | 120.50 ±16.26 |
| *Sex* |  |  |
| Female | 47 | 113.36 ±17.49 |
| Male | 58 | 114.05 ±10.52 |
| Unknown | 1 | 107.00 |
| *Marital status* |  |  |
| Single | 74 | 114.65 ± 11.00 |
| Married | 28 | 115.04 ±8.93 |
| Divorced | 1 | 104.00 |
| Widowed | 0 | N/A |
| Unknown | 3 | 113.67 ±7.37 |
| *District of residence* |  |  |
| 3 | 21 | 115.48 ±11.98 |
| 5 | 24 | 113.33 ±10.74 |
| 9 | 31 | 114.45 ±8.59 |
| 13 | 24 | 114.13 ±10.67 |
| 17 | 1 | 125.00 |
| 19 | 1 | 127.00 |
| Unknown | 4 | 116.50 ±13.23 |
| *Education* |  |  |
| Completed primary | 31 | 112.52 ±9.67 |
| Completed secondary | 61 | 114.44 ±10.76 |
| At college / university | 7 | 123.00 ± 5.39 |
| Bachelor's degree | 2 | 119.50 ±10.61 |
| Postgraduate degree | 0 | N/A |
| Other | 2 | 119.50 ±17.68 |
| Unknown | 3 | 114.00 ±7.94 |
| *Occupation* |  |  |
| Employed fulltime | 10 | 117.30 ±7.51 |
| Employed part time | 13 | 116.77 ±10.92 |
| Unemployed | 59 | 113.71 ±10.89 |
| Student | 23 | 113.43 ±8.24 |
| Unknown | 1 | 141.00 |
| *Proximity to someone with a mental illness* |  |  |
| Yes | 64 | 115.59 ±10.16 |
| No | 39 | 112.85 ±10.71 |
| Not sure | 2 | 121.50 ±4.95 |
| Unknown | 1 | 108.00 |

### Key findings from thematic analysis of focus group discussions

From the eleven total study participants recruited for the focus group series, only seven were able to participate in one or more sessions **(Table 4)**. The four who were not able to participate had planned to join the focus groups virtually but were unable to participate due to unstable internet connection caused by network issues in Malawi.

Despite those challenges, the seven participants engaged proactively in all sessions, providing their views and critique of the MHLq, and proposed changes on how to make it more applicable to the cultural context of Malawi. Thematic analysis of transcripts from focus group discussions suggested a more pessimistic view on the state of mental health literacy for young people in Malawi than the quantitative MHLq results suggest. The overall sentiment was that knowledge and understanding of what mental health is, along with the causes and different types of mental disorders was very low in Malawi. Focus group participants attributed this discrepancy between results of the MHLq responses and their own personal experiences and perspectives to three key factors: 1) epistemic exclusion, 2) conceptual misfits, and 3) translation challenges **(Figure 1)**.

**Figure 1.**
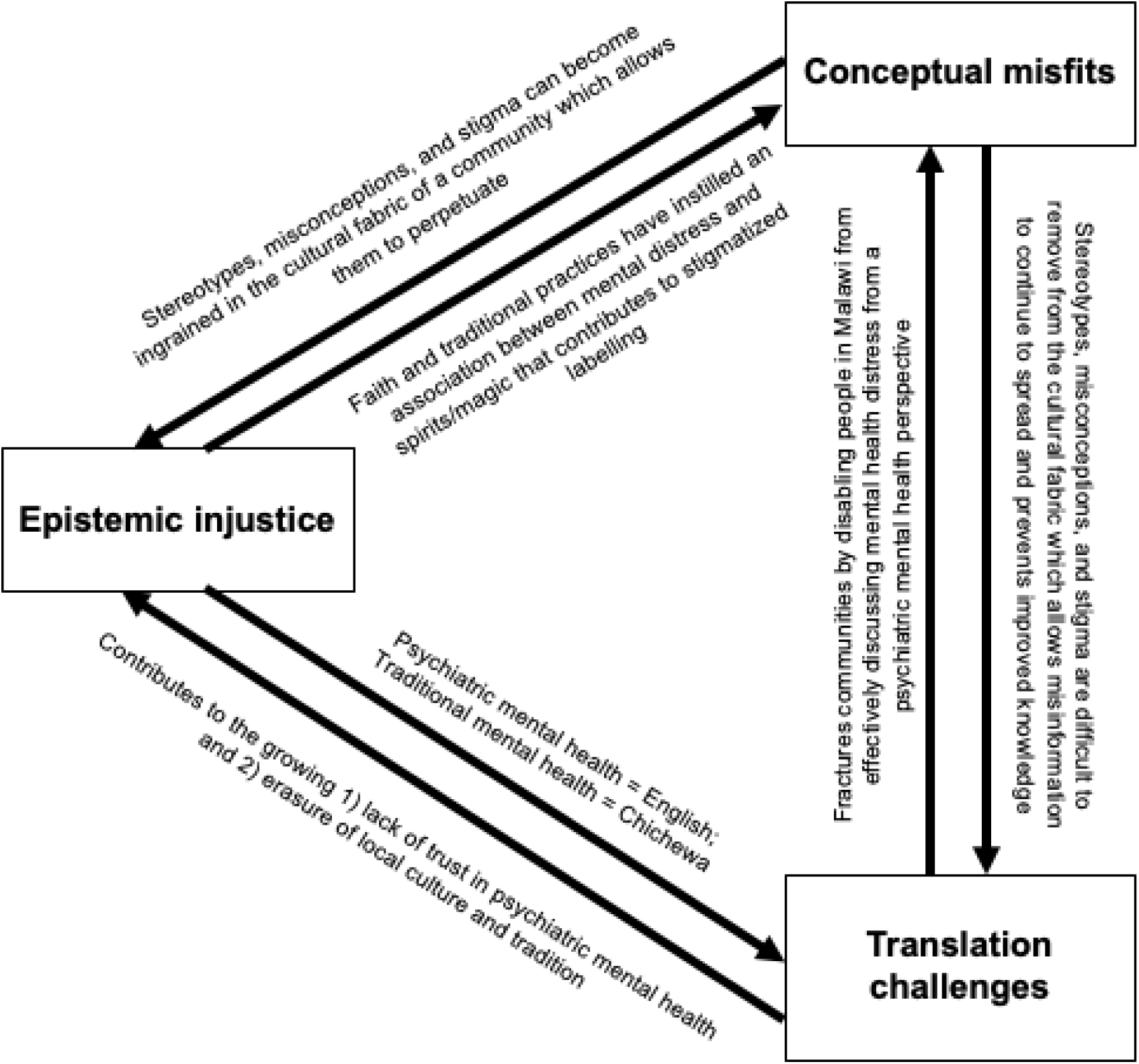
Thematic map of focus group discussions. The figure depicts how these themes are linked in a thematic map.

### Theme 1: Epistemic exclusion

One key factor that focus group participants felt contributed to the discrepancy between the MHLq results and their own personal experiences with mental healthcare in Malawi was that the social and cultural context in Malawi was missing from the questionnaire items. This was exemplified through two key aspects of Malawian culture that study participants identified as missing: a) community and b) faith.

### **a)** Community

Malawian culture is rich in its own traditions and social norms which focus group discussions revealed were not represented in the MHLq. Specifically, one individual noted the importance of community and social cohesion in Malawi and how the MHLq did not reflect this.

> *“I think from the Malawi context…they should have included more about [how] we are very socially connected…Your relationships, like your family and friends, are very important in Malawi. For example…even that question about…doing something enjoyable, maybe they could have asked [about] spending time or being with your close family and friends or [how the] people you love contribute to good mental [health] because I think a lot of people in Malawi, when they’re in trouble, they lean a lot on…their family, their friends, those people in [their] community…I think…that could be added as a self-help [strategy].”*

The other study participants agreed with this observation and advocated for the inclusion of this aspect of daily life into the MHLq items.

> *“[Having] someone to talk to [is important in Malawi], especially if you are going through a situation like a break up or something. So…[there] should be something [about] social [connection].”*

Some participants (n=2) felt that the strong sense of social cohesion within their communities would encourage them to share feelings of mental distress.

> *“I would tell my friends…because they would understand if I were to tell them…what I am going through. It [would be] a relief like, oh, my problem is gone because I’ve shared it with someone.”*

Nevertheless, other participants (n=4) did not feel comfortable sharing feelings of mental distress due to fear of a negative reaction.

> *“If I had a mental disorder…I wouldn’t seek my friend’s help. I wouldn’t seek my parents’ help. I would just go straight to the professional and explain to them what I’m going through. Because…especially here in our country…I feel people would be prejudiced, they would judge.”*

Study participants revealed that this discomfort in sharing feelings of mental distress is related to their general belief that: *“Here in Africa, we’re not as sensitive to how people feel. There’s a lack of sensitivity of how someone feels.”*

> *“There are people who are silently suffering from depression, which is a mental illness, and people just regard it as [weakness]…Toughen up, it happens, it’s normal, it’s part of life…Yet, people are killing themselves over it.”*

### **b)** Faith

Another important aspect of Malawian culture that does not appear in the MHLq items is faith and religion. Many people in Malawi reliably attend religious services which is connected to Malawi’s strong sense of community due to the vast social network religion provides (Yeatman & Trinitapoli, 2008; Kendall, 2019). This was corroborated by study participants who highlighted the importance of faith in Malawian culture, both socially and personally.

> *“Going to church, it’s not just a social activity it’s…faith…And faith has its own characteristics that…gives hope.”*

Religion also plays a role in Malawian culture through traditional mental health practices. Traditionally, people in Malawi seek the help of spiritual healers and religious leaders to address mental distress (Steinforth, 2017).

> *“Some people, their perception of mental illness is very superstitious, so they might end up having conclusions that this individual has demons [or] supernatural forces that are [causing] this behaviour. So, when they go to such spiritual healers and religious figureheads, they end up wanting assistance to take away those supernatural forces.”*

> *“[Spiritual healers and religious leaders] act as frontline practitioners…They take up that slot, before going to the psychologist or the psychiatrist.”*

However, study participants noted a generational shift in the importance of faith, where young people have tended to stray away from religious beliefs and practices more so than previous generations. This led participants to show scepticism towards the benefits of spiritual healers and religious leaders which reflects the anecdotal shift in culture among young people in Malawi away from faith and religion.

> *“Don’t take someone with mental illness to a theologian. It’s not going to help. They need a psychologist. That’s it.”*

In summary, focus groups sessions revealed epistemic exclusion in the MHLq as it did not measure attitudes towards community and faith, which participants identified as key aspects of Malawian culture. Despite agreeing on the importance of these two cultural concepts, participants disagreed on their impact on mental health. While some felt that their own communities were important for maintaining good mental wellbeing, others felt that they were unable to discuss mental health with their families and friends due to fear of stigma. Also, while some viewed faith and religion as an anchor for hope and socialisation, others stated that the role of the church in traditional healing hinders people from seeking mental health support. The MHLq’s inability to draw out these cultural epistemologies reveals a weakness the tool has for reliably measuring mental health literacy for young people in Malawi.

### Theme 2: Conceptual misfits

Participants reflected on how the aspects of Malawian culture discussed in Theme 1 may contribute to widespread mental health stigma that persists due to common stereotypes and misconceptions.

> *“It’s just the inability… to understand what is going on with the person. So, let’s say [friend’s name] …started hallucinating. We’ll say … that maybe she had ‘chamba’ or…some evil spirits are with her and we’ll come up with a lot of names and…labelling…And at the end of the day, it’s never something nice.”*

‘Chamba’ is the Chichewan word for cannabis which many people in Malawi believe causes mental health issues. Participants suggested that this stigma towards mental illness stems from the stereotype that *“all crazy people are the same.”*

> *“[People in Malawi believe] that…all mental illnesses are the same. People don’t categorise them. He has depression, they crazy. Anxiety, they crazy. Bipolar, they just crazy.”*

Participants indicated that this occurs due to misconceptions among people in Malawi about the causes of mental illnesses. As a consequence, people often blame either supernatural forces or the individual experiencing mental ill-health.

> *“In Malawi, it’s either witchcraft or substance abuse…such as marijuana…If [people] abuse marijuana and then have a mental illness… [people] will [think] the illness is from that. So, then it’s okay, we understand. But if…you [do not] have any physical proof…that’s the stereotype…here.”*

> *“If the reason for your mental problems isn’t a curse, people in Malawi usually say it’s your fault because they link it to things like you drink too much or you’re abusing substances. So, you’re just being reckless…it’s actually your fault, so we don’t need to waste our time or money on you.”*

Overall, focus group sessions revealed that the MHLq was not able to accurately depict the mental health stereotypes, misconceptions, and stigma that participants stated exist in Malawi due to conceptual misfits between the stereotypes and misconceptions in the MHLq items and those that young Malawians experience. Participants identified a need for an explanation for someone’s behaviour to be able to understand it in Malawi, including evil spirts or drug use. The MHLq does not include any items that measure beliefs regarding evil spirits. It did, however, include one item that measures beliefs regarding drug use (Item #24: Drug addiction may cause mental disorders) which 100 respondents (93.5%) either agreed or strongly agreed with. This suggests that there may be a need for more items on the causes of mental health issues that are conceptually relevant to the local context in Malawi.

### Theme 3: Translation challenges

Lastly, study participants discussed translation challenges when talking about mental distress in Malawi throughout focus group sessions.

The relatively high mean total mental health literacy score ((114.63) compared to the Portuguese study (105.27)) suggests that MHLq respondents in rural Malawi are knowledgeable about mental health. However, study participants disagreed, remarking that although MHLq respondents may have been aware of disease entities like schizophrenia, they were very likely uncertain about what they were. Study participants felt that this gap in knowledge was common among young people in Malawi.

> *“People witness people who have schizophrenia. But because of the lack of mental [health] literacy, I don’t even think they can differentiate it…The majority of Malawians are just going to put it in one word, ‘wapenga’ or ‘ndi wa misala.’”* In English, ‘wapenga’ translates to ‘you are crazy’ and ‘ndi masala’ translates to ‘they are mad/insane.’ Study participants agreed that neither of these Chichewa terms can be translated as schizophrenia as a disease entity and perceived both terms to be stigmatizing.

Surprisingly, the MHLq data revealed that 82 respondents (76.6%) either agreed or strongly agreed with item# 3: *“People with schizophrenia usually have delusions (e.g. they may believe they are constantly being followed and observed).”* However, one participant, who was involved in coordinating fieldworkers for the MHLq data collection, provided a potential explanation for this discrepancy.

> *“I think there was bias on that [item] because some people asked, what is [schizophrenia]? … and I had to just give the signs and the symptoms. So, I think they just used that interpretation to answer …because there was no [translation for schizophrenia] in [the] Chichewa version. Or other [respondents] skipped this [item].”*

Focus group participants explored this further, stating that there is a lack of non-stigmatizing words in the Chichewa language to talk about mental distress or an absence of direct translation for specific mental health terms. This corroborates similar findings from a previous study (Jumbe et al., 2022). For example, in the Chichewa version of the MHLq, there is no translation for schizophrenia provided. When asked if they knew of a Chichewa translation, participants agreed that no translation existed and *“there would just be a description of something close to schizophrenia.”* Because of this lack of language to define some mental disorders in Chichewa, some participants (n=4) felt it would be difficult to discuss their own mental health with friends and family.

> *“[If] you tell somebody…here in Malawi… ‘I have schizophrenia,’… [they’ll say], ‘what’s that?’ And then in Chichewa, you’d have to say, ‘ndili ndi misala.’ And they’re just going to say, ‘what, you’re crazy?’…They don’t understand that it’s an illness. You’re sick, not crazy. So, even if you say “ndikudwala malungo a ubongo,” it sounds like you’re crazy.”*

Additionally, participants indicated that while people in Malawi may have heard the terms psychologist and psychiatrist, it is very unlikely that they know what these roles are and how they are different. This is emphasised by the lack of direct translation for either term in the Chichewa translation. General lack of knowledge about psychiatrists and psychologists is reflected anecdotally by the participant who was coordinating fieldworkers for MHLq responses.

> *“I was looking for a way to explain what a psychiatrist does. So, I would tell them [that they] …help people [with] mental issues, those who will give drugs to people*. *So, people actually have a sense that, okay, it’s a doctor for mental disorders. [However], the only problem in Malawi is explaining who a psychologist was because…people actually suggested someone who reads their minds by just looking at [them].”*

These discussions suggest bias in the MHLq responses, with a high likelihood that some respondents were using guesswork based on information provided by the surveying fieldworkers. One participant suggested that this could be overcome by including an “I don’t know” option to the Likert scale response options.

> *“The answer you give is…biased because you would just maybe say I neither agree nor disagree, instead of being honest that you don’t know.”*

Furthermore, there is no direct translation for psychologist and psychiatrist. The Chichewa version of the MHLq uses descriptions for these terms in item# 5, 8, 17, and 29. Study participants back-translated these descriptions into English:

Psychologist translation: *“someone who is an expert of the brain or the mind”*

Psychiatrist translation: *“somebody who is an expert in craziness [or] crazy people”*

Participants all agreed that these types of descriptions were stigmatizing and needed to be changed to improve knowledge and understanding of mental health. In prior research, the concept of the idioms of distress, alternative ways to express distress related to cultural meaning, has attempted to include culturally relevant and appropriate terms for discussing psychiatry in non-Western settings by merging local understandings with psychiatric disorders (Nichter, 1981). Although this concept has been criticised for conflating traditional discourses with psychiatric disease entities, it is also a powerful tool for making psychiatry accessible to non-English speaking people (Patel, 1995; Nichter, 2010). During focus group discussions, one participant suggested how this could be done with schizophrenia.

> *“I think one other solution might be borrowing the word by abbreviating it…Instead of schizophrenia, you just say ‘shizofenia’ in Chichewa.”*

Focus group sessions revealed that MHLq scores may have been inflated by the absence of direct translation of psychiatric terms into Chichewa. Psychiatric terms in the Chichewa version of the MHLq were written in English due to this absence of direct translation, which study participants confirmed. They, however, proposed suggestions for how to overcome this barrier through the addition of an ‘I don’t know’ response to the Likert scale and adaptations to the English words to be more acceptable to Chichewa speakers (e.g. ‘shizofrenia’ instead of schizophrenia).

## Discussion

### Main findings

This study contributes to the limited body of mental health research in Malawi by investigating the applicability of a mental health literacy tool using perspectives and lived experiences of young people to adapt this tool to the local epistemology or “truths”. This is a novel approach to assessing mental health literacy tools, as it employs a material semiotics theoretical approach by interrogating what the MHLq does and how it impacts both the psychiatric mental health field and young people in Malawi (Yates-Doerr, 2020). This does not deny the materiality of the MHLq or its importance as a metric for assessing mental health literacy, but rather it uncovers its meaning (Yates-Doerr, 2020). Uncovering the meaning of the MHLq in the local context revealed epistemic injustice in the prioritisation of psychiatric mental health “truths” over Malawi’s traditional and cultural “truths.” For the MHLq to become cultural applicability and better represent the state of mental health literacy among young people in Malawi, decolonising strategies should be operated to adapt the questionnaire.

All focus group participants raised concern that the quantitative results from the MHLq did not accurately depict mental health literacy among young people in Malawi. They felt the questionnaire neglected to include key aspects of Malawian culture and life (e.g. community and faith) which have both positive and negative impacts on mental health. Although the MHLq discussed stereotypes and erroneous beliefs, it was not able to capture the specific ways that participants felt stigma persists and prevents mental health literacy from improving in Malawi. This was illuminated by the connection between an apparent lack of mental health knowledge and non-stigmatizing language available in Chichewa, which highlighted barriers to learning about and understanding mental distress. Overall, participants felt that more culturally appropriate MHLq items and the addition of an “I don’t know” option on the Likert scale were necessary to make the MHLq more applicable for young people in Malawi, and ultimately, produce more reliable results.

The relatively high mean total mental health literacy score from MHLq respondents in rural Malawi, compared to the score from respondents in the Portuguese MHLq study (Dias et al.,2018), was unexpected because Malawi has comparatively fewer mental health resources that generally result in lower mental health literacy rates (Kutcher et al., 2015; Kutcher, Wei, & Coniglio, 2016). However, there is conflicting evidence in the current literature about whether increased knowledge about mental health reduces stigma (Thornicroft et al., 2007). Therefore, quantifying mental health literacy, using a metric tool like the MHLq may not fully capture the complexity of mental health understanding in Malawi. This demonstrates a general weakness in questionnaires as they tend to operate under one specific “truth” which may not account for alternative epistemologies (Yates-Doerr, 2020; Law, 2015).

### Recommendations for adaptations to the MHLq

From the results of this study, it is recommended that the MHLq be adapted to operate the “truths” of both psychiatric and traditional mental health to generate results that are both psychiatrically relevant and valid but also appropriate and meaningful for young people in Malawi. It is suggested that this be done following the aspects of decolonisation set out by de Sousa Santos that are relevant to this study: 1) recovering diverse knowledge systems and 2) decolonizing research and history (de Sousa Santos, 2014).

### 1) Recovering diverse knowledge systems

Study participants identified that community and faith are two aspects of life that are ingrained in Malawian culture. However, many of the MHLq items were framed from an individualistic perspective (e.g. Item# 26: Doing something enjoyable contributes to good mental health). Although “something enjoyable” could include spending time with family or going to church, the item’s framing suggests something that an individual does for their own mental health, rather than a collective activity within a community. This individualistic framing is often associated with cultures in high-income countries, where psychiatric mental health knowledge and concepts originated, and therefore, may influence the way that young Malawians interpret the items. Creating new or rewriting existing items in Factor 4: self-help strategies that investigate the relationship between mental health and community and faith as cultural factors would strengthen the accuracy and impact of the responses. Adding these elements would improve the reliability of this questionnaire because current literature tends to understate the role that culture has on stigma (Koschorke et al., 2016).

Study participants also suggested that many of the stereotypes and erroneous beliefs described in the items from Factor 2: erroneous beliefs or stereotypes were also associated with those from high-income countries. This could be improved by including some of the erroneous beliefs that study participants identified as common among their peers. For example, the belief that *‘all crazy people are the same’* could be framed in the MHLq as ‘All people experiencing mental distress are crazy’ or ‘All people experiencing mental distress should be rejected by their communities.’ This is important because the aim of using the MHLq and other similar tools is to understand baseline mental health literacy in order to inform mental health education. Therefore, if the questionnaire is testing the stereotypes of young people from another culture, the education will also address these stereotypes and ignore those prevalent in the local setting. This allows the locally specific beliefs to perpetuate without targeted intervention.

Further, study participants highlighted that psychiatric terms come from the English language and do not have a direct translation in Chichewa. MHLq respondents likely relied on guesswork or support from the field workers who disseminated the survey, which may have influenced the results for Factor 1: knowledge of mental health problems. This could be overcome by incorporating local epistemologies into psychiatric mental health to offer more meaning to clinical terms. One participant suggested creating Chichewa words from the English ones (e.g. schizophrenia could become *‘shizofenia’*). However, creating new idioms would be outside of the scope of a cultural adaptation to a questionnaire. In the absence of local idioms that accurately represent the psychiatric terms used, the most appropriate adaptation for this factor would be to add an ‘I don’t know’ option to the Likert scale, as one participant suggested. Items should also be added to the MHLq to understand attitudes towards mental health from a traditional mental health perspective (e.g. *‘misala’*). These options offer a more appropriate meaning to the MHLq for young people in Malawi without rejecting the materiality of psychiatric mental health. This is a step towards decolonising mental health literacy by incorporating traditional mental health “truths” alongside psychiatric mental health “truths;” thus, acknowledging the value of each for young people in Malawi.

Study participants had similar reflections regarding the items from Factor 3: first aid skills and help-seeking behaviours. Out of the six items in this factor, four are statements about seeking help from a psychologist or psychiatrist. Study participants indicated that these are not terms that young Malawians are typically familiar with and do not have a direct translation in Chichewa. While local idioms and an ‘I don’t know’ option on the Likert scale would help to improve the accuracy of responses, it is also recommended that the MHLq include items that measure attitudes towards traditional healers and religious leaders to understand young people’s relationship with these figures as mental health support. In addition, items that discuss mental health professionals more generally may be more suitable as psychiatric mental health resources are scarce in Malawi and there has been a greater focus on community mental health workers to help meet demand, close the treatment, and prevent mental disorders (Jervase et al., 2022). While this includes the “truths” of traditional mental health, it also does not deny the “truths” of psychiatric mental health, and is therefore, appropriate and meaningful to both epistemologies.

### 2) Decolonising research and history

This research highlights the importance of involving the people directly affected by your work in the work itself. Without insights from study participants, the pitfalls of this MHLq would remain largely unknown, making it difficult to culturally adapt the questionnaire for young people in Malawi. Consulting with young people in Malawi is a first step to improving mental health literacy in a way that is both culturally appropriate and clinically relevant. Future research should aim to co-produce mental health literacy and education tools to ensure that this balance is achieved from the beginning and to build community ownership.

### Strengths and limitations

Study strengths included the use of a mixed methods approach for deeper understanding of mental health literacy, investigating quantitative MHLq responses while also exploring how and why respondents might have arrived at their answers through qualitative focus group discussions. Using online platforms for communication (Twitter, WhatsApp, and Microsoft Outlook) and focus group sessions (Zoom), while offering in-person options was also a strength. This both removed the geographical barrier and overcame some network issues in Malawi due to digital poverty. Ultimately, these varying options for conducting research open future opportunities for more cross-cultural work. The use of focus groups to assess the applicability of such mental health literacy questionnaires is novel, providing essential contextual and linguistic insights that may not otherwise be considered. This signifies the importance of working towards epistemic justice and acknowledging all of the “truths” that exist within a given context.

Nevertheless, quantitative results were limited by disparities in the number of MHLq respondents within each demographic category, which may have contributed to the lack of significant differences between groups. The MHLq responses were also limited by potential response bias, as the MHLq is a self-reported questionnaire. Further, digital poverty (network issues) were also a limitation, despite the preventative measures applied, with some off-campus participants unable to join all focus group sessions consistently. Additionally, the sampling method was biased towards recruiting focus group participants who were either university students or graduates, English speaking, and interested in mental health. This excluded representation from the majority of Malawians who are primary school-educated, non-English speaking, and reside in rural areas (UNESCO, 2019; OECD, 2018). Another limitation of this study is that its results lack generalisability due to its specific aim to improve the MHLq. That said, the general suggestions to include locally relevant idioms and cultural practices and to co-production in future research are relevant for other mental health literacy tools.

## Conclusion

The study aimed to assess the applicability of a mental health literacy tool for measuring mental health literacy among young people in Malawi. MHLq responses resulted in unexpectedly high mental health literacy scores which revealed a possible bias in the responses due to a lack of cultural applicability. Focus groups dissected the MHLq, illuminating on attitudes and knowledge regarding mental health among young people in Malawi and the effect this may have had on responses to MHLq items. Mental health literacy tools should avoid relying on self-help and help-seeking strategies and stereotypes from high-income countries. Instead, they should seek to focus on what resonates with the local population. They should also avoid using terms that are unfamiliar to the cohort they are assessing, and instead, opt for local idioms where appropriate or allowing the opportunity for participants to state that they do not know the answer. This should be developed further through future research into how to incorporate psychiatric mental health terms into the local language, building upon the concepts of idiom of distress. Lastly, mental health literacy tools should be created, tested, and implemented alongside the local population to ensure both cultural and psychiatric applicability. These recommendations would benefit future research by offering appropriate tools to assess mental health knowledge and awareness, which can more accurately and effectively inform mental health education for young people in Malawi.

## Data Availability

All data produced in the present work are contained in the manuscript

